# Deep learning-based localization of bounded edentulous spaces in intraoral occlusal images

**DOI:** 10.1101/2025.06.24.25330144

**Authors:** Ehsan Shirdel, Saeideh Azizipour, Mary Magdalyanova

## Abstract

**Objectives:** This study developed a deep learning pipeline to localize bounded edentulous spaces with missing teeth FDI number directly from intraoral photographs, eliminating reliance on radiographs or controlled imaging.

**Materials and Methods:** A total of 4,373 intraoral occlusal images were included in the study. Of these, 367 images with at least one bounded edentulous space were held out for pipeline evaluation. The remaining 4,006 images were split into training (80%), validation (10%), and test (10%) sets for model development. The pipeline included ResNet-18, YOLOv8m and ResNet-101. Bounded edentulous were inferred based on FDI discontinuities and localized via spatial interpolation.

**Results:** The pipeline was evaluated on a test set of 367 occlusal images, comprising 4,964 expected teeth. This equated to a precision of 88.2%, recall of 89.1%, an F1-score of 88.6%, and MCC of 0.875. The pipeline showed stable diagnostic performance for actual use cases and made full inference in 9 seconds per image on consumer-grade hardware, indicating its viability for use with standard clinical workflows.

**Conclusions:** The pipeline showed reliable ability to detect and localize bounded edentulous spaces. Its consistent performance across a wide range of different clinical cases justifies its use as a diagnostic tool in routine dental practice.

**Clinical Significance:** Our results show that accurate detection of edentulous spaces is now possible even from unstandardized clinical images and is less dependent on radiographs and manual charting. This simplifies workflows in day-to-day practice, aids precision in orthodontic and implant planning, and makes AI tools more accessible for use in those practices with minimal image infrastructure.

## 1. Introduction

Bounded edentulism is a very common clinical situation with important functional, aesthetic, and psychological implications for the patients [1]. Patients with missing teeth suffer from impaired chewing, modified phonetics, occlusal discord, and decreased quality of life [2, 3]. Proper identification and recording of edentulism is, therefore, important for optimal treatment planning in prosthodontics, implantology, and orthodontics [3].

Traditional methods of edentulism identification mainly use the panoramic radiograph and manual notation, and there are important clinical drawbacks to each of them. Radiographic imaging is impractical in many resource-poor situations because it requires exposure to ionizing radiation and access to specialty equipment [4]. In addition, the task of dental charting is often relegated to auxiliary staff, including dental assistants, who do not receive the diagnostic training needed to perform the task with consistency and data fidelity [5]. As the use of non-specialist opinion adds variability and increases the chances of documentation error, diagnostic fidelity is finally jeopardized.

Even with the increased use of digital photography in dental workflows, intraoral occlusal photographs are still underexploited in the context of automated diagnostics [6]. The reason is partly because they are characterized by variability in angulation, illumination, interference from the soft tissues and the presence of artifacts like restorations and orthodontics [7]. However, the images provide highly defined, radiation-free visualizations of occlusal anatomical structures that are amenable to robust artificial intelligence (AI)-based decision-making if exploited appropriately [6, 7].

The majority of currently available AI-based dental research has focused mostly on radiographic datasets obtained from controlled, standardized acquisition protocols [6]. Although the systems enable algorithm development to take place, they disproportionately exclude the heterogeneity, variability, and unpredictability of the actual clinical settings [8, 9]. Consequently, the resultant models tend to possess limited translational potential in general dental practices, outreach programs, and resource-scarce settings—the settings where radiographic infrastructure is either not available, cost prohibitive, or contraindicated [6, 8, 9]. This discrepancy highlights a critical lack of applicability of existing AI systems to everyday, chairside diagnostic processes that utilize freely available, non-radiographic modalities like intraoral occlusal photographs [10].

In response to this shortfall, this work introduces a deep learning-based pipeline that is able to detect and localize bounded edentulous space directly from intraoral occlusal photographs. The approach doesn’t use radiographic inputs nor controlled acquisition protocols and is intended to run under real-world clinical variability. The developed pipeline not only provides a new framework but also addresses a critical clinical necessity for automatic dental charting with diagnostic-level precision.

In situations where thorough patient records are not available or manual data is susceptible to inaccuracy this system provides a consistent alternative. Using routine intraoral occlusal photographs for bounded edentulous space detection and localization, it enhances diagnostic accuracy and supports the inclusion of AI in scaled dental informatics processes.

## 2. Materials and methods

### 2.1 Ethics

Ethical approval for this study was obtained from the Ethics Committee of Sechenov University (The approval number will be added to the manuscript following the final decision by the authors and the completion of the committee’s internal review prior to formal publication), and the study was conducted in accordance with the Declaration of Helsinki. Prior to participation, all patients provided written informed consent authorizing the use of their anonymized intraoral photographs for research purposes. For participants under the age of 18, informed consent was obtained from a legally authorized guardian, who also accompanied the minor during image acquisition. Consent forms containing identifiable information were securely stored. All image data used in this study were fully anonymized prior to analysis. In compliance with data protection regulations, no personally identifiable information was collected, stored, or disclosed by the research team.

### 2.2 Dataset

#### 2.2.1 Patient selection

Participants consisted of patients aged 12 years and above, with no retained primary teeth at the time of photography. The cohort included patients from diverse ethnic backgrounds and exhibited a wide range of socioeconomic and educational profiles, reflecting the urban population. No exclusion criteria were applied based on gender, ethnicity, income level, or prior dental history, unless the patient or their legal guardian was unwilling to participate in the study.

#### 2.2.2 Dataset allocation

A total of 4,764 intraoral occlusal photographs—both the maxillary and mandibular arch images—were gathered. All the images had been taken under standardized clinical conditions with a Canon EOS 800D DSLR (Canon Inc., Tokyo, Japan) camera and a standard 18–55 mm lens with LED ring flash for consistent occlusal light and anatomical accuracy among the various cases. Adobe Photoshop CC (Adobe Inc., San Jose, CA, USA) software had been used for standardized cropping. Each image was resized to a consistent resolution of 2000 × 2803 pixels. All images were exported as high-quality JPG to retain structural information like morphology of the cusps, fissure patterns, and interproximal contours.

A total of 391 images with the presence of at least one third molar, either partially erupted or fully erupted, in the occlusal area were excluded. The teeth were excluded from the analysis based on their low occurrence in the dataset and could lead to distributional instability and bias during training of the model [11]. Clinically, third molars often don’t fully erupt because of the anatomical constraints and the lack of space. Moreover, in contemporary dental practice, prophylactic or therapeutic extraction of third molars is routinely performed, rendering their representation inconsistent and their inclusion less relevant to standardized occlusal analysis [12, 13].

Of the remaining 4,373 images, 367 cases exhibiting at least one single-tooth bounded edentulous space were deliberately withheld from all model training and validation processes. This was done to ensure that during the final evaluation of the full pipeline, the models had no prior exposure to these images. This ensures enabling a reliable assessment of missing-tooth localization performance. The remaining 4,006 images (2,004 maxillary and 2,002 mandibular) were used to train and evaluate classification models, and were split into training (∼80%, n=3,206), validation (∼10%, n=400; 200 maxillary, 200 mandibular) and test (∼10%, n=400; 200 maxillary, 200 mandibular) subsets.

#### 2.2.3 Annotation protocol

All annotation procedures were performed manually by a team of six specialist dentists, each with a minimum of five years of clinical experience. The annotators were divided into two groups of three. Initially, each group independently annotated a subset. Upon completion, the annotated datasets were exchanged between the two groups for cross-review and verification. All annotators were fully informed of the purpose of each annotation stage and were trained in the operational definitions and classification criteria relevant to the task.

Maxillary and mandibular images were first separated into two distinct groups. A binary label was then assigned to each image: class 0 for maxillary and 1 for mandibular. All images were annotated using makesense.ai and Roboflow. A bounding box was drawn around each visible tooth, and each tooth was initially classified into one of seven anatomical classes based on its morphology.

After annotation, a full copy of the dataset was generated, resulting in two identical sets of images and annotations, each prepared to support a distinct modeling objective. In the first dataset, intended for the Residual Neural Network-based classifiers (ResNet-101), each image was duplicated, and all visible teeth were individually cropped based on their bounding boxes. The cropped tooth images were then organized into 14 folders corresponding to 7 anatomical classes for the maxillary arch and 7 for the mandibular arch. In the second dataset, which retained the full occlusal images, all detailed anatomical class labels were replaced with a single unified label. This simplified labeling was applied to train the medium version of the You Only Look Once model (YOLOv8m) as a single-class object detector focused solely on detecting and localizing teeth, regardless of their type or position.

### 2.3 Models and training

These models were trained using Google Colab Pro+, with an NVIDIA A100 GPU. A staged training strategy was employed to avoid propagation of errors between stages and enable customized optimization for each model based on its particular task. The design supported easier performance monitoring and debugging at every stage of the pipeline.

#### 2.3.1 Jaw classification

A ResNet-18 convolutional neural network in PyTorch 2.5.1 with Compute Unified Device Architecture (CUDA 12.4) compatibility was utilized to predict occlusal images as either maxillary or mandibular. The model architecture was trained from scratch with random weights, avoiding the dependence on pre-learned features from natural image datasets (e.g., ImageNet), potentially introducing semantic bias or domain shift. The last fully connected (FC) layer was substituted with a two-unit output layer followed by a dropout layer (P = 0.5) to reduce the risk of overfitting.

All images were resized to 224×224 pixel and normalized to standard ImageNet mean and standard deviation (mean = [0.485, 0.456, 0.406]; std = [0.229, 0.224, 0.225]) for compatibility with the model architecture. Instead of using standard real-time data augmentation during training, controlled offline data augmentation was used pre-model initialization.

For each jaw class, the dataset was split randomly into four equal parts (25% each): group A had the original unrotated images; group B had images rotated 90° clockwise; group C had images rotated 90° counter-clockwise; and group D had images rotated 180°. This pre-defined strategy of data augmentation was chosen with the intent of inducing orientation-invariant feature learning, where the model could learn to detect clinically important soft tissues and anatomical cues—like the palatal rugae or the position of the tongue—without being distracted by image orientation [14].

A SoftMax activation came with the use of the Cross-Entropy Loss method, which internally includes SoftMax and negative log-likelihood computations. Accordingly, the output of the final layer of the model produced raw logits to enable the stable computation of gradients data loading was performed through the *torch*. *utils*. *data*. *Dataloader* with a batch size of 64 and two worker processes for parallel loading. The AdamW optimizer (weight decay = 1e-5, learning rate = 0.001), appropriate for deep learning with sparse updates and adaptive rates, was used for training. Training proceeded for a maximum of 300 epochs with patience-based early stopping (patience = 30 epochs) based upon validation accuracy [16]. ResNet-18 is selected for its good balance between model capacity and computational efficiency, with enough representational ability for the anatomical classification as a binary task with minimal overfitting danger to moderately sized medical datasets [15].

#### 2.3.2 Tooth detection

For localizing the individual teeth within occlusal images, an object detection model based on YOLOv8m (Ultralytics v8.3.134) was trained in single-class mode. All bounding boxes were put into a single class and optimized to identify their locations regardless of anatomical type or quadrant. The model was trained for 300 epochs with an image resolution of 640×640 pixels, a batch size of 32, and AdamW optimizer (learning rate = 0.001, weight decay = 1e-5). A dropout rate of 0.4 was implemented to counter overfitting [16].

Model validation took place after every single epoch using mean Average Precision (mAP) at Intersection over Union (IoU) 0.5, which acted as the main assessment metric. An early stopping strategy was implemented: training was automatically terminated if no improvement in mAP@50 was observed over 40 consecutive epochs. The model checkpoint achieving the highest mAP@50 across training was saved as the final version for deployment. As YOLOv8m internally uses a sigmoid activation function per bounding box in single-class mode, no explicit SoftMax was applied. The confidence output for each detection is a direct representation of a tooth’s presence probability within the estimated region.

The YOLOv8m model version was chosen to balance representational ability and computationally tractability optimally, which suits it to identify small, morphologically congruent elements like teeth from intraoral occlusal images. Larger models (e.g., YOLOv8l/x), though delivering slight improvements in accuracy, have much greater memory and inference expenses, and are suboptimal for clinical or resource-limited deployment. Lightweight versions (YOLO8n/s), in turn, tend to be limited by lower depth of features and receptive field extent, causing lower precision in localizing highly spaced dental elements [17]. YOLOv8m offers an intermediate solution that balances deeper backbone encodings and improved spatial resolution preservation and lasts longer even with generalization under high-resolution dental imagery, without becoming inefficient at training [18].

#### 2.3.3 Anatomical tooth classification

Two separate deep convolutional models based on ResNet-101 were trained to classify cropped individual teeth into seven anatomical categories, one for the maxillary arch and one for the mandibular arch. Both models were identically configured and differed only in the anatomical source of their respective datasets. Each network was initialized with pretrained weights from ImageNet to take advantage of low-level feature generalization, and the FC layer was replaced by a seven-unit linear classifier.

The images for inputs are resized to 224×224 pixels and normalized via standard channel-wise ImageNet statistics: mean = [0.485, 0.456, 0.406] and standard deviation = [0.229, 0.224, 0.225] for red, green, and blue channels, respectively. Geometrical and color-based augmentation is not used. The ResNet-101 models were trained based on the PyTorch deep learning platform version 2.6.0 with support for version 12.4 of CUDA and using an AdamW optimizer, which had a learning rate of 1×10⁻⁴ and weight decay of 1×10⁻⁵. This has been demonstrated to enable deeper network convergence and improved generalization, especially when paired with batch normalization. Training was done using a batch size of 64 for a maximum of 300 epochs. An early stopping mechanism was employed, monitoring epoch-level validation accuracy as the performance criterion. If validation accuracy did not increase for 50 epochs continuously, training stopped to prevent overfitting and wasteful computation [16]. This prevented model capacity from becoming over-invested beyond where generalization becomes significant.

The dropout had been intentionally omitted from ResNet-101 models since the architecture already has built-in strong regularization by virtue of the residual connections and batch normalization between each convolutional block. Since stable gradient flow, depth of the network, and access to a very large and diverse dataset exist, further stochastic regularization would not be needed [19].

Empirical validation revealed that adding dropout (p = 0.4) before the final classification layer lowered validation accuracy from 0.95 to 0.93 for the maxillary model and from 0.94 to 0.91 for the mandibular model, which reflects a negative effect on generalization. ResNet-101 was chosen due to its added architectural depth (101 convolutional layers), and high representational capacity (approx. 44.5 million parameters), which support elaborate hierarchical feature extraction. This is especially helpful when discriminating morphologically similar dental classes where differences in shape and boundaries are paramount [20, 21].

### 2.4 Inference pipeline

(Fig. 1) The fully trained system then processes each occlusal photograph through a 9-step pipeline sequentially to output tooth-level anatomical classifications, numbering by FDI World Dental Federation (FDI), and missing tooth localization. The inference process goes through the following sequence:

a. **Input image:** The image is supplied as input.
b. **Jaw classification:** Jaw Classification: The photograph is initially passed through ResNet-18 classifier to decide if it’s the maxillary (class =0) or mandibular (class=1) arch.
c. **Tooth detection:** The image is processed by YOLOv8m to detect all the visible teeth, and bounding box coordinates are output for each detection.
d. **Tooth cropping:** A bounding box is employed to crop out each tooth region from each original image. The cropped patches are later resized and normalized according to the input specifications of the anatomical classifiers.
e. **Anatomical class assignment:** Each identified tooth was cropped from the bounding box which had been predicted by YOLOv8m and resized to 224×224 pixels. Normalized standard channel-wise ImageNet preprocessing was carried out before classification. Depending upon the jaw type identified from step 2, each cropped tooth is forwarded to the respective ResNet-101 model (maxilla or mandible) to identify its anatomical class (1– 7). The model produced one of seven classes of anatomy: central incisor, lateral incisor, canine, first premolar, second premolar, first molar, or second molar. Class prediction was obtained through argmax by applying it to the vector of SoftMax outputs.

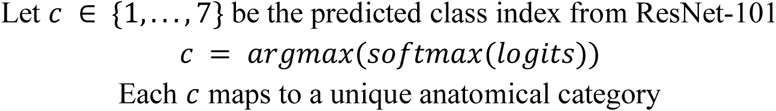
f. **Image reconstruction:** Predicted bounding boxes and class numbers were overlaid to recreate an annotated image. Each box was marked with the tooth class number.
g. **FDI Number mapping:** A logic-based mapping function was employed to allocate a complete FDI number to each tooth. This function merged anatomical class prediction (*c*), normalized horizontal x-center coordinate (*x_c_*), and jaw label prediction (*j*). The quadrant number (*q*) and final FDI number were calculated based upon jaw and lateral position.

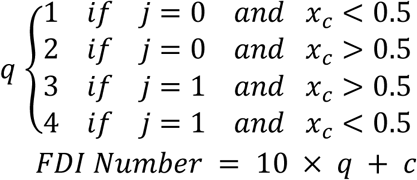
h. **FDI Sequence analysis:** The set of predicted FDI numbers was first sorted and deduplicated. To identify missing teeth, expected FDI sequences for each jaw were used as references. Missing labels were inferred by detecting gaps between adjacent numbers in the sorted sequence. A minimum of four valid FDI detections was required per image to proceed with interpolation.

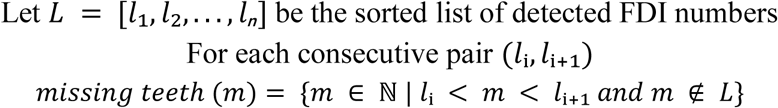
i. **Spline-based interpolation:** To estimate the position of missing teeth, univariate spline functions were fit to the x- and y-coordinates of the detected teeth as a function of FDI number. The splines captured the curvature of the dental arch and allowed spatial interpolation of tooth positions. Only second-order (*k* = 2) splines with no smoothing (*s* = 0) were used.

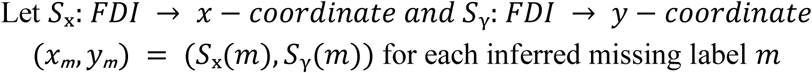
j. **Edentulous localization:** Each interpolated (*x*, *y*) coordinate was transformed into pixel space using the original image dimensions. A square bounding box with side length equal to 6% of the image’s minimum dimension was drawn around the inferred position. The label ′*Missing*: [*FDI*]’ was placed above the box using OpenCV’s text rendering functions.

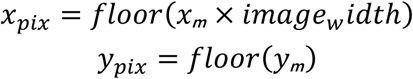

**Fig. 1.**
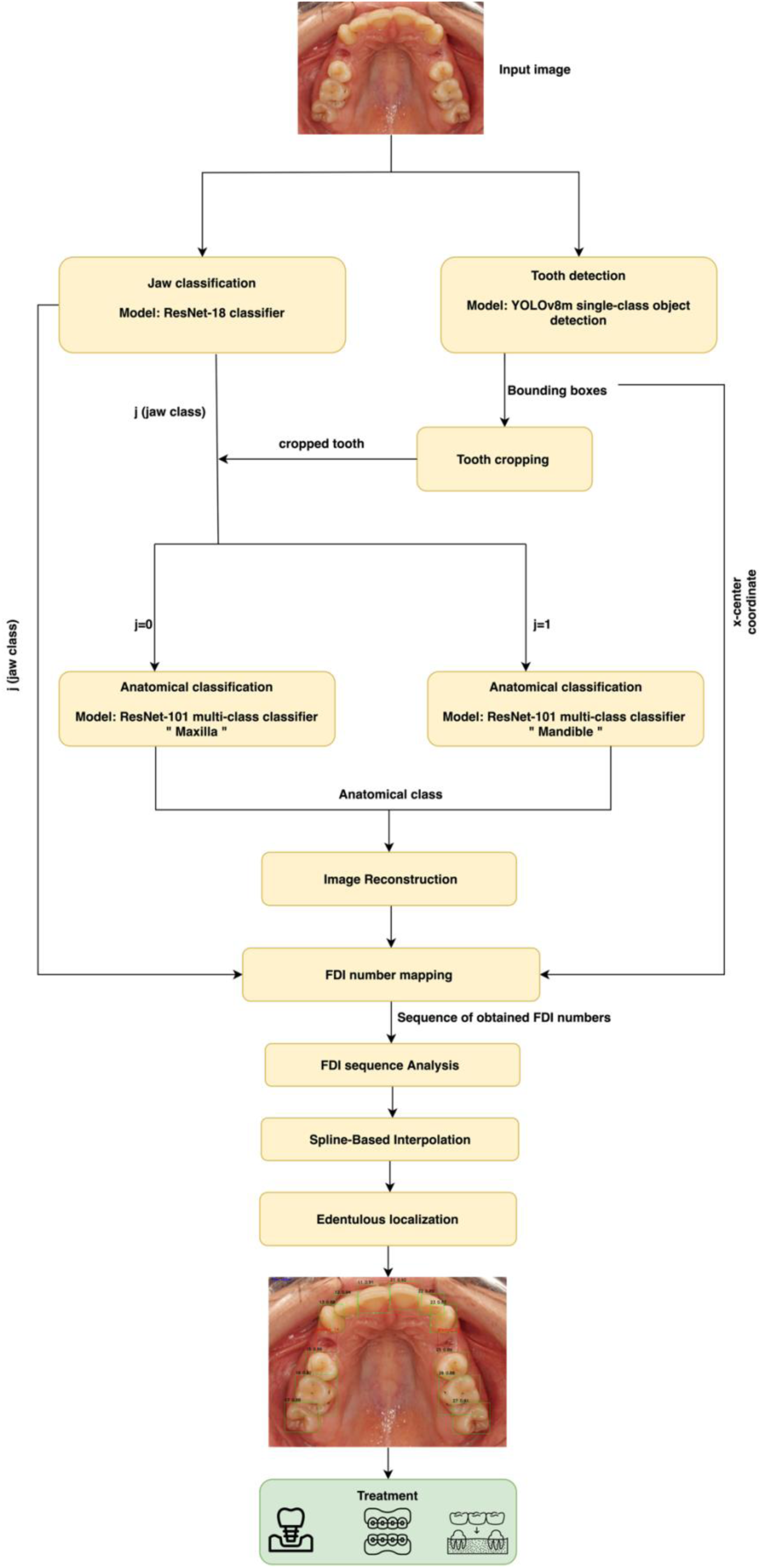
Overview of the proposed pipeline for bounded edentulous detection.

## 3. Results

### 3.1 Performance of individual deep learning modules

Each model underwent independent assessment using a test dataset of 400 annotated occlusal images. Performance measures were calculated individually to verify each module before integration.

#### 3.1.1 Jaw classification (ResNet-18)

The model had an overall accuracy of 0.91, which is indicative of consistent binary discrimination between classes of jaws. To test class-specific performance, precision, recall, and F1-score were calculated per class. For Class 0 (maxillary images), these had values of 0.93, 0.89, and 0.91, respectively. For Class 1 (mandibular images), their respective values were 0.89, 0.93, and 0.91. These balanced scores indicate consistent performance across both anatomical categories.

(Fig. 2a) Grad-CAM visualizations revealed that the model did not focus on the curvature of the dental arch when distinguishing between maxillary and mandibular occlusal views. Instead, it consistently attended to posterior intraoral regions, particularly detecting the presence or absence of the tongue or lingual frenum whereas in maxillary views, this region appeared anatomically absent or recessed [22].

**Fig. 2.**
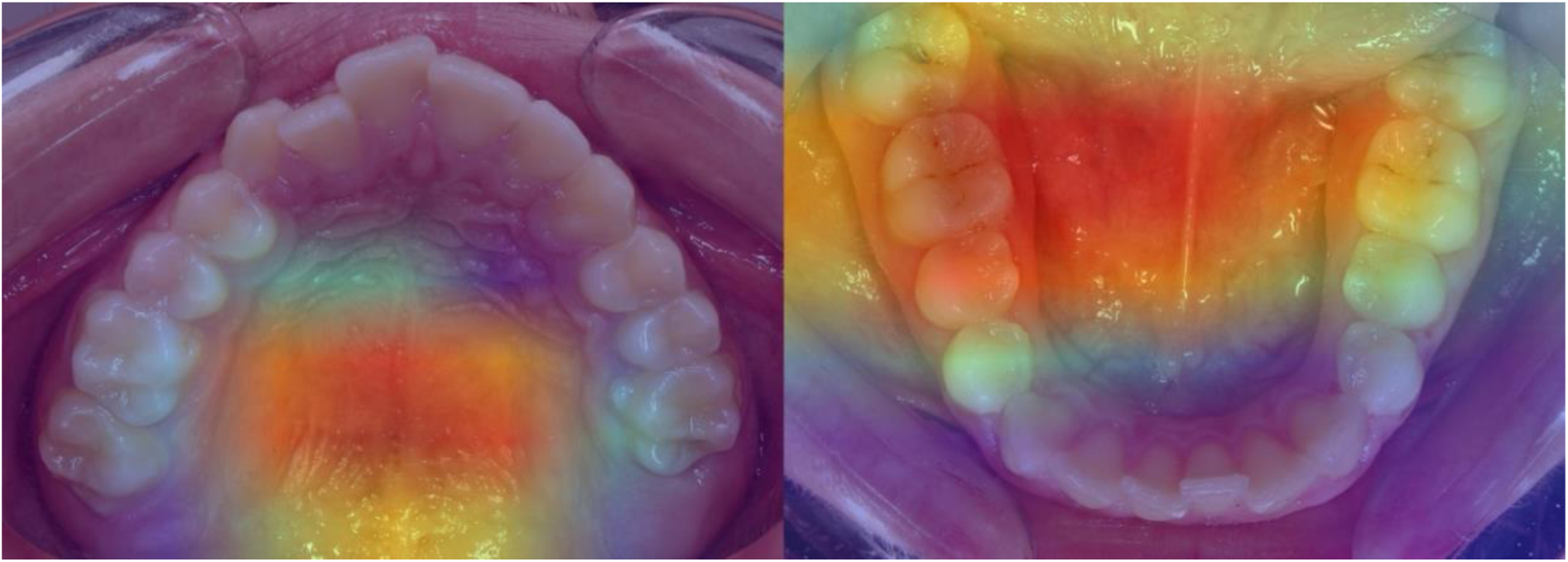
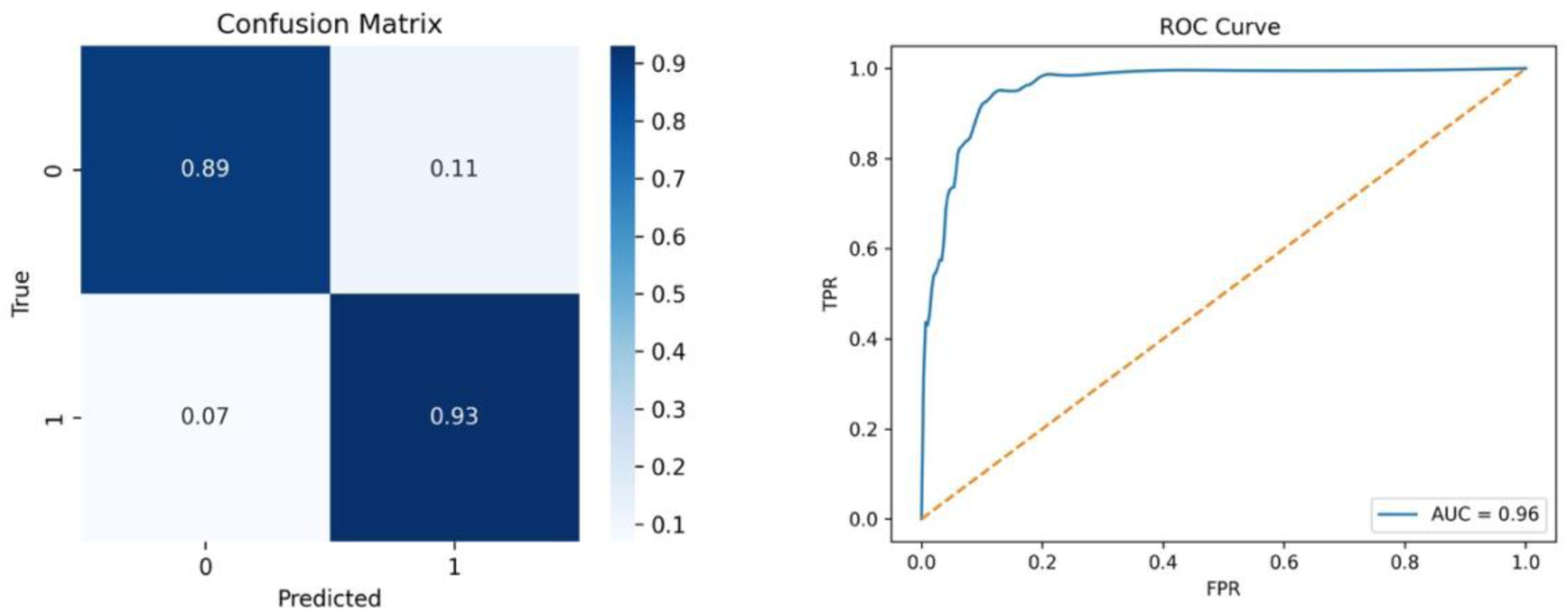
a. Grad-CAM visualizations for ResNet-18 showing the regions of attention in maxillary (left) and mandibular (right) images. Warmer areas indicate regions the model focused on most, while cooler areas reflect regions with minimal contribution to the prediction. b. Confusion matrix and ROC curve of the ResNet-18 (AUC = 0.96). Class weights (maxillary: 2.0; mandibular: 1.0) were used to address data imbalance and reduce classification bias.

(Fig. 2b) The confusion matrix illustrates an excellent classification performance of the ResNet-18 model for the binary task of jaw type. The model has correctly identified 89% of maxillary and 93% of mandibular samples, indicating high sensitivity for both classes. Rates of misclassification were relatively modest, with 11% of maxilla and 7% of mandible predictions assigned to the wrong class. (Fig. 2b) The shape of the ROC curve further supports the model’s ability to discriminate. The curve has a steep slope toward the top-left corner, which indicates a high rate of true positives even for relatively low false positive thresholds. The Area Under the Curve (AUC) of 0.96 indicates excellent separability between classes, indicating that feature representations learnt by this model are well-suited for this classification problem.

#### 3.1.2 Tooth detection (YOLOv8m)

Precision, recall, and F1-score were computed based on predicted bounding boxes with a confidence ≥ 0.8. A detection was classified as a true positive if its IoU with any ground truth box exceeded 0.5; otherwise, it was considered a false positive. Since the dataset was single-class, all metrics reflect detection performance rather than inter-class discrimination. The model achieving a mAP@0.5 of 0.9867, mAP@0.5-0.95 of 0.7558, precision of 0.9736, recall of 0.9675, and an F1-score of 0.9866. The overall accuracy was 0.9736.

(Fig. 3) The histogram of IoU distribution shows how accurately the bounding box predictions are spatially related to ground truth. All predictions which appear in the plot had confidence and IoU ≥ 0.80. The distribution occupies values between 0.88 and 0.91, which shows a significant overlap for most of the predictions. That the density around these values is very sharp indicates that the model localizes object boundaries consistently with very little variance [23].

**Fig. 3.**
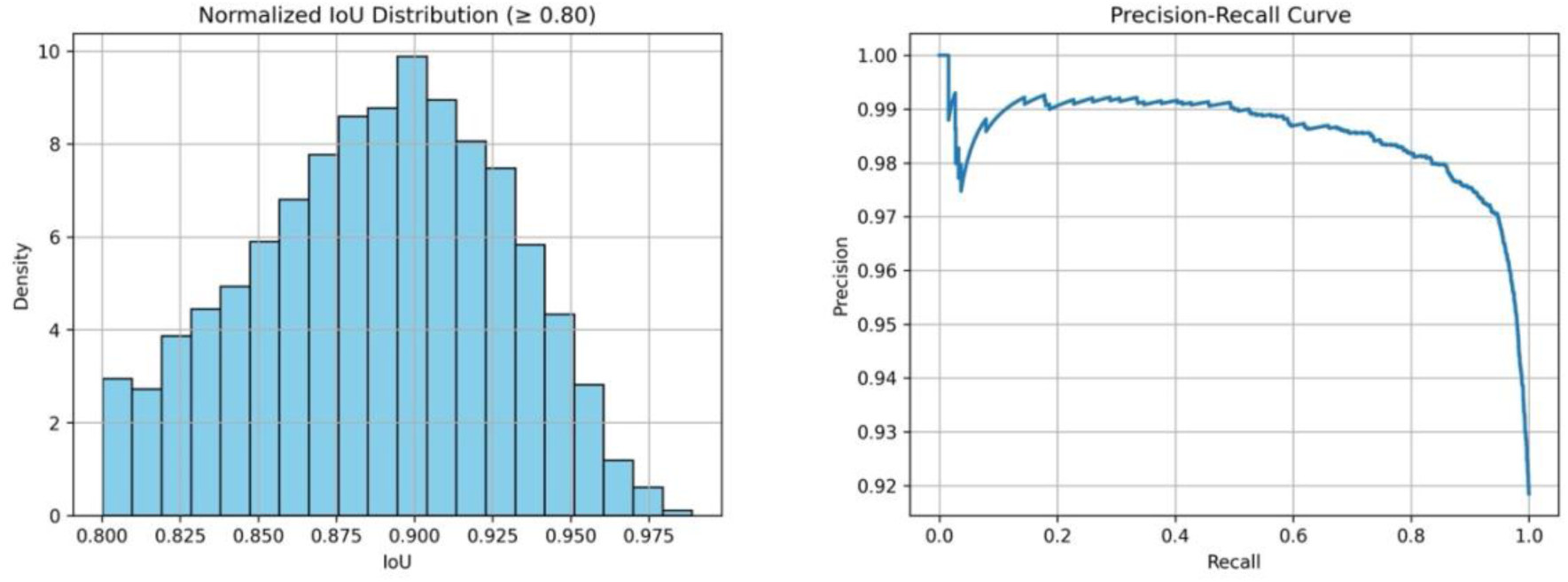
IoU histogram and precision-recall curve of YOLOv8m. An initial drop in precision is observed due to limited predictions at low recall, followed by consistent high precision (∼0.98) across most thresholds, with a final drop near maximum recall

(Fig. 3) The Precision–Recall curve shows stable performance spanning a very wide range of recall levels. Precision is stable and high (≥ 0.97) for nearly the entire range of recall, and decreases very little as recall nears 1.0. Such behavior is consistent with well-calibrated confidence thresholding and minimal false positive rate. The smooth curve and absence of extreme fluctuations are an indication of model reliability across different detection sensitivities.

#### 3.1.3 Anatomical classification (ResNet-101)

The ResNet-101 models evaluated for maxilla and mandible achieved overall accuracies of 0.95 and 0.94, respectively. To assess class-wise performance, precision, recall, and F1-score were computed separately for each of the seven anatomical tooth classes in both arches (Table 1).

**Table 1.** Class-wise precision, recall, F1-score and support for anatomical classification using ResNet-101.

| Class | Maxilla |  |  |  | Mandible |  |  |  |
| --- | --- | --- | --- | --- | --- | --- | --- | --- |
|  | Precision | Recall | F1-score | Support | Precision | Recall | F1-score | Support |
| Central incisor (1) | 0.98 | 0.93 | 0.96 | 400 | 0.92 | 0.96 | 0.94 | 399 |
| Lateral incisor (2) | 0.87 | 0.99 | 0.93 | 400 | 0.93 | 0.91 | 0.92 | 400 |
| Canine (3) | 0.99 | 0.92 | 0.96 | 390 | 0.97 | 0.95 | 0.96 | 369 |
| First premolar (4) | 0.92 | 0.97 | 0.94 | 348 | 0.92 | 0.95 | 0.93 | 393 |
| Second premolar (5) | 0.95 | 0.93 | 0.94 | 400 | 0.97 | 0.93 | 0.95 | 378 |
| First molar (6) | 0.98 | 0.96 | 0.97 | 400 | 0.94 | 0.98 | 0.96 | 393 |
| Second molar (7) | 0.97 | 0.96 | 0.97 | 348 | 0.99 | 0.93 | 0.95 | 321 |

Given the multiclass nature of the tooth classification task the use of accuracy alone is insufficient to characterize the model’s discriminative ability across all categories [26]. There might be classes of teeth that are intrinsically easier to distinguish based on their individual shape and prevalence in the database. Others might be more variable and overlap and therefore tend to be prone to misclassifications. Such fine-grained analysis enables detection of which anatomical classes are especially prone to false positives (expressed by low precision), as well as false negatives (expressed by low recall), and allows detection of imbalances in performance caused by class imbalance or by anatomical similarity [24, 25].

(Fig. 4a) Furthermore, it enhances clinical interpretability by ensuring that diagnostically significant misclassifications are not obscured by the aggregation of performance into a single overall accuracy metric. (Fig. 4b) the model attained uniformly high normalized accuracies for all seven maxillary classes, and class-wise distinguishability was corroborated by t-SNE projections. Class 2 attained Acceptable accuracy and formed a dense, isolated cluster in t-SNE space, suggesting highly discriminative morphological attributes and little intra-class variability. Class 1 (0.93) and Class 3 (0.92), by contrast, had medium-level confusion with Class 2 (6% and 7%, respectively), which can be explained by their proximate anatomical locations within the anterior segment of the maxilla, where central and lateral incisors have overlapping crown shape, especially when viewed occlusally.

**Fig. 4.**
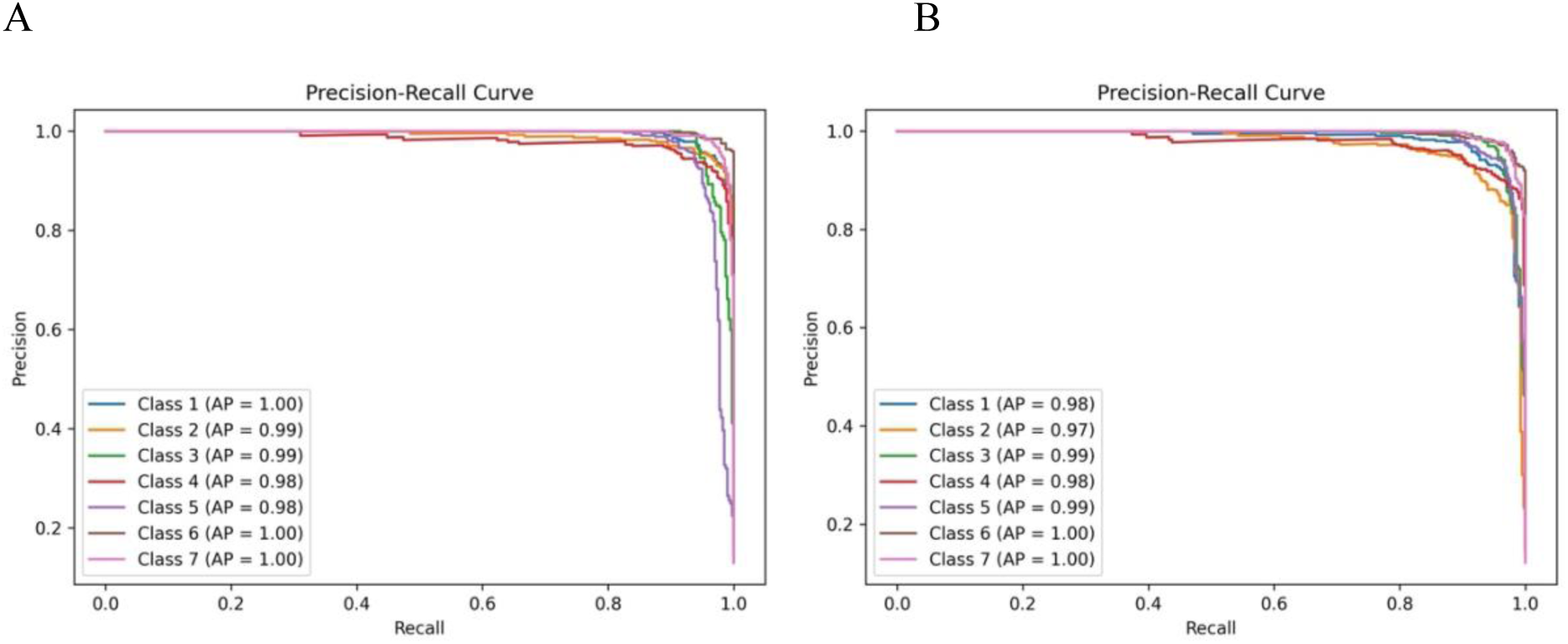
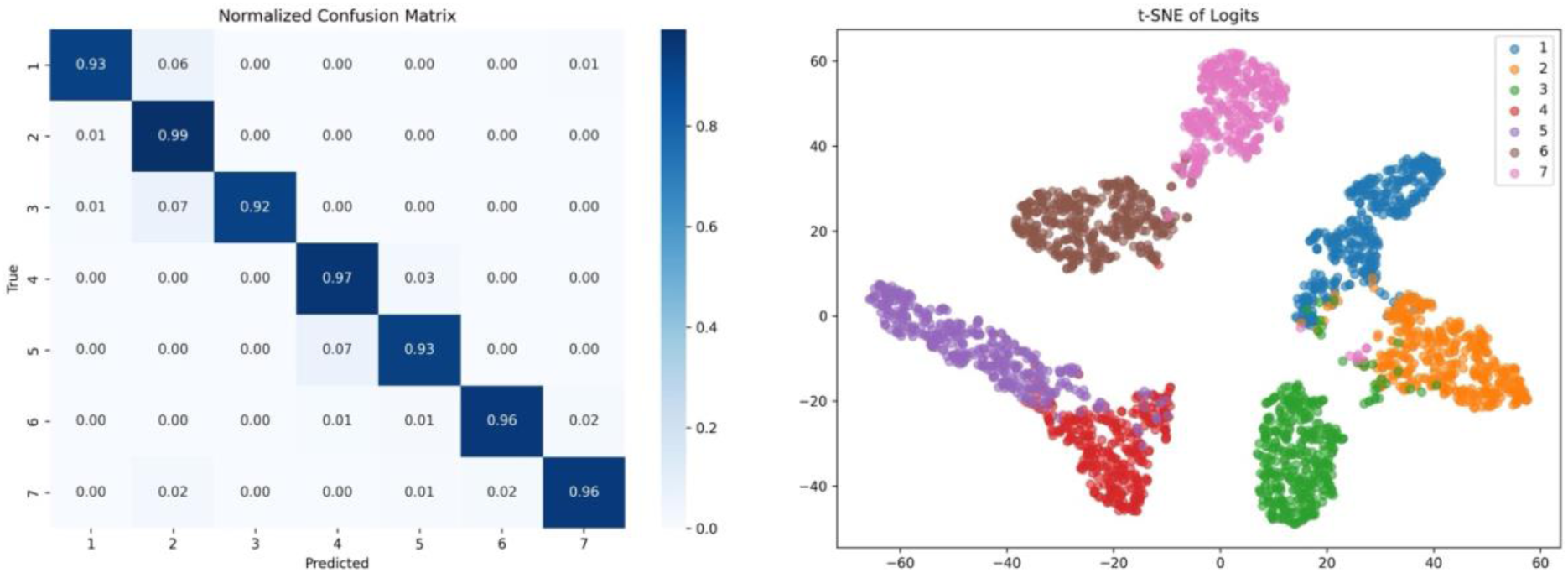
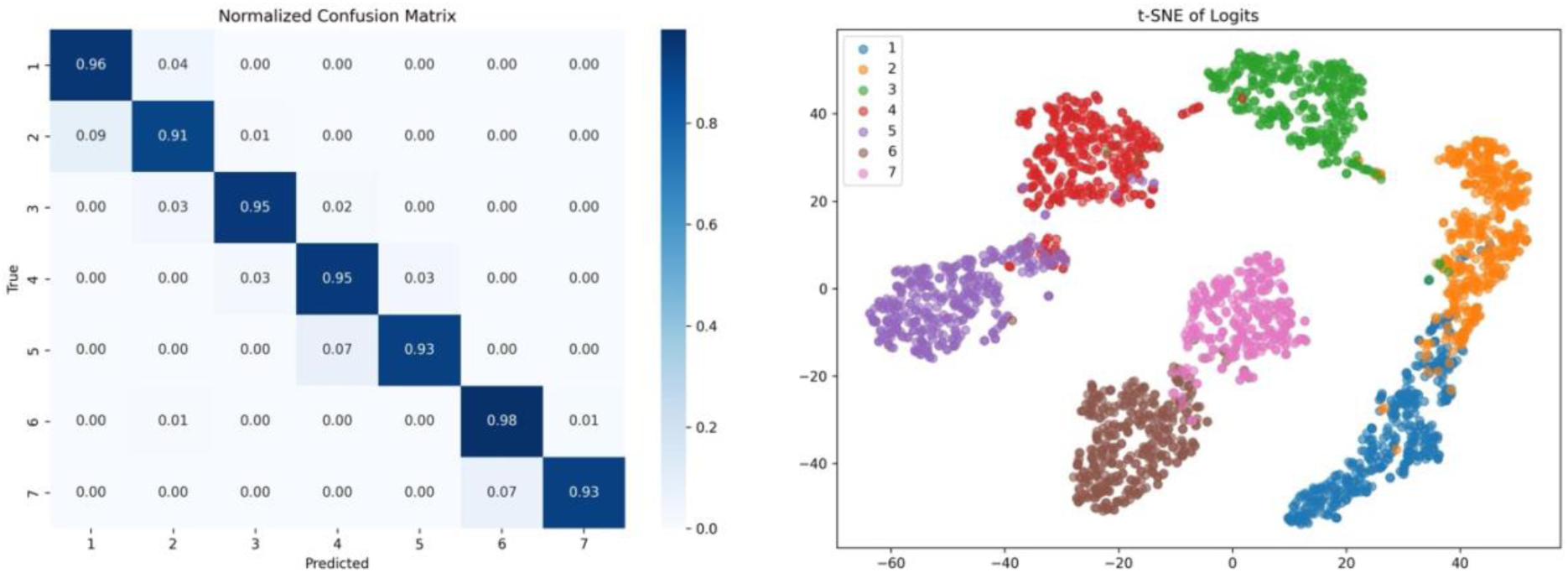
a. Precision-recall curves for ResNet-101 classification of maxillary (A) and mandibular (B) teeth. Drops in precision indicate false positives. b. Normalized confusion matrix and t-SNE plot for maxillary ResNet-101. Class weights of 1.2 and 1.5 were applied to Class 2 and Class 5, respectively, to address class-specific imbalance. c. Confusion matrix and t-SNE plot for mandibular ResNet-101. Most errors occur between neighboring classes. No class weighting was applied during training.

Classes 4 and 5, which are first and second premolars, had reciprocally incorrect classification (3% and 7%, respectively), and partial cluster contiguity within t-SNE space; such behavior is natural considering their analogous cusp morphology and contour outline, which are less discernible in 2D representations. Classes 6 and 7 (molars) had good accuracy (0.96 each), but minor confusion between each other and Class 2 most likely stems from occlusal angle distortions or overlapping shadows impacting feature detection. Overall, incorrect classifications are consistent with clinically reasonable patterns of anatomical similarity and visual ambiguity.

(Fig. 4c) ResNet-101 demonstrated excellent mandibular classification, with errors matching anatomical and imaging-driven uncertainty. Class 6 (0.98) has highest accuracy, displaying a dense and compact cluster in the t-SNE plot, consistent with large occlusal surface area and singular molar shape that allows for discriminative feature learning. Class 1 has excellent performance (0.96) but displays a 4% confusion for Class 2, which might be caused by visual contiguity of anterior mandibular teeth; central and lateral incisors have subtle shape differences. Class 2 has lowest accuracy (0.91), displaying a 9% confusion for Class 1, consistent with challenging separability of tightly contacted anterior teeth due to minimal crown-height variability.

Classes 3 and 4 exhibit confusions despite their excellent overall accuracies (0.95 each), likely caused by mandibular canine transitional shape and 2D view similarity to premolars. Class 5 has 0.93 accuracy and 7% confusion for Class 4, consistent with their adjacent anatomical location and similar cusp patterns of premolars. Class 7, similar to Class 5, has 0.93 accuracy and mild confusions into Class 6 (7%), though their t-SNE groups are largely separable. Spatial disposition of t-SNE groups, particularly elongation and occasional interpenetration between Classes 1–2 and 3–5, confirms that most confusions stem not from model failure but from intrinsic shape gradients and limitations of image capture specific to mandibular occlusal surfaces.

### 3.2 End-to-end evaluation of the complete AI pipeline

(Fig. 5) The full pipeline was evaluated on a reserved test set of 367 images. 4964 teeth were expected, of which 443 were clinically missing and 4521 were present. A true positive (TP) was defined as a case in which the pipeline correctly identified a missing tooth by accurately assigning its FDI number and detecting its absence at the appropriate anatomical location within the dental arch. A true negative (TN) was recorded when the pipeline correctly recognized a present tooth, assigned the correct FDI label, and did not erroneously report a missing space in that region. A false positive (FP) occurred when the pipeline incorrectly predicted a gap in place of a present tooth—typically due to FDI mislabeling or misinterpretation of inter-dental spacing. Conversely, A false negative (FN) described an actual missing tooth which the pipeline did not detect, either missing the area completely or by assigning its FDI identifier to an adjacent existing tooth.

**Fig. 5.**
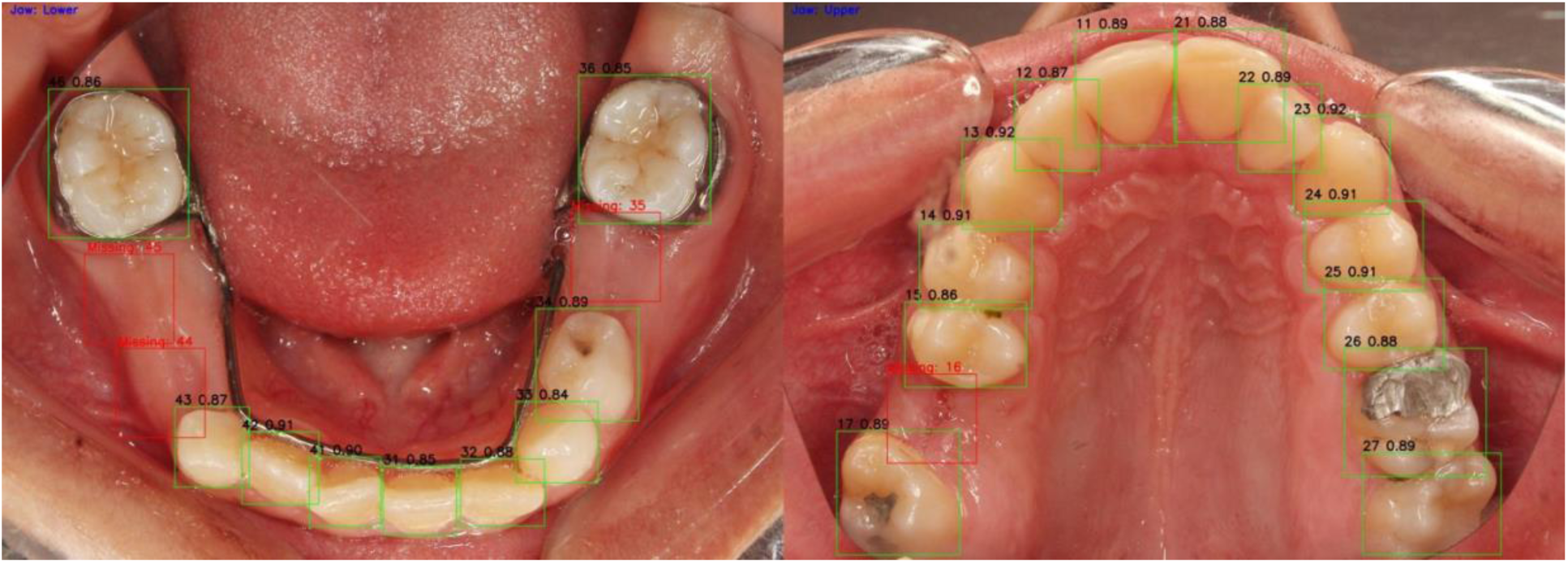
Result of end-to-end pipeline. Bounded edentulous space detected despite obstruction (left). Despite the presence of amalgam, remaining cusps of tooth 26 were clearly recognized.

These definitions were chosen to capture clinical and anatomical dental arch intricacies wherein detection validity depends not just upon the missing tooth’s identity (through FDI numbering), but upon correct topographic context as well. Unlike straightforward binary judgments insensitive to labeling errors, such a design takes into account clinically significant cases wherein a tooth is assigned an incorrect number—potentially leading to false assumptions about edentulous spaces. By mandating that a true positive be correct numerically (FDI) and spatially (within the right place within the arch), the assessment guarantees consistency with clinical standards of diagnosis.

The final predictions of the pipeline were independently reviewed by three board-certified dental specialists to determine the clinical correctness of missing tooth identification. Based on expert adjudication, the pipeline correctly identified 404 missing teeth (TP) and 4457 present teeth (TN). It failed to detect 49 missing teeth (FN) and erroneously predicted 54 false gaps (FP). These results correspond to a precision of 0.882, a recall of 0.891, a F1-score of 0.886 and a Matthews Correlation Coefficient (MCC) of 0.875.

In clinically relevant classification tasks particularly those involving a disproportion between negative and positive cases conventional metrics such as accuracy can be fundamentally misleading [26, 27]. This study focuses on identifying bounded edentulous, a minority class from the dataset, wherein most labels map to existing teeth. In imbalanced situations like these, classification algorithms can yield spuriously high-test scores by intentionally skewing their outcomes towards majority classes and underperforming for minority class representations of greater diagnostic significance [27]. To counter such a bias induced by imbalance, we took the MCC as our first-choice metric. MCC provides a statistically grounded assessment by considering all four values of the confusion matrix: true positives, true negatives, false positives, and false negatives [28]. Unlike such measures, which get biased towards one class or another and hence can yield spuriously good scores, MCC is sensitive to both classes of classification mistake and therefore gives a truer reflection of actual diagnostic utility [28, 29].

Most importantly, MCC accords with clinical reasoning: missing teeth are not merely uncommon but also are of central importance to treatment planning, and their omission or their false detection can lead to significant downstream mistakes [28, 29, 30]. By reflecting both the model’s capacity to detect actual edentulous regions and its restraint in avoiding false positives, MCC delivers a more balanced and trustworthy measure of performance than accuracy or F1-score alone [28]. To evaluate clinical applicability, the full pipeline was run on a standard 2.0 GHz quad-core Intel Core i5 system with integrated Intel Iris Plus Graphics. End-to-end inference per image averaged 9 seconds, supporting its integration into real-time, chairside diagnostic settings without specialized hardware.

## 4. Discussion

To maximize clinical generalizability of the proposed pipeline, no images were excluded from training or evaluation, except those containing third molars. The universal inclusion of images ensured that model representation space contained a broad range of anatomical and clinical variability present in real-world images from intraoral photography.

The ResNet-18 model had more common cases of misclassifications in images from the maxilla during tasks, owing predominantly to orthodontic appliances like palatal expanders, nance buttons, or transpalatal arches. The metallic devices pose non-biological features that conceal native palatal morphology and compromise spatial patterns generally employed by convolutional levels for discrimination. The model suffered from intrinsic limitations to segregate such non-anatomic artifacts from relevant anatomical cues and thus decreased robustness in orthodontically treated maxillary cases.

The YOLOv8m model had a main limitation caused by excessive pooling of saliva, especially from posterior mandibular areas, where mucosal or glandular specular reflections created contrast-heavy noise that confused the detector. Detection errors often occurred when there were orthodontic brackets, intraoral jewelry, gold crowns, and big amalgam restorations, which distorted the natural shape and visual texture of the teeth and caused bounding box errors and detection failures. More difficulties appeared when there were restorations of big amalgam fillings spanning over two-thirds of the crown surface, these restorations concealed the natural tooth shape and added considerable variability, which disrupted the model’s generalizability.

Imaging domain-specific augmentations, adaptive exposure control during capture, or post-processing stages to suppress artifacts from regions of high reflection might be needed to enhance robustness. A larger dataset and more variability during clinical capture conditions could in addition provide greater generalizability.

ResNet-101 experienced persistent deficiencies when there are maxillary images where morphological similarity between premolars resulted in false classifications. Nonstandard canine positions, like partial eruption or mesial drifting, tended to cause it to be mistaken for the lateral incisors when diminished by angulation or occlusal distortion. Metallic orthodontic brackets also added artificial edges to confuse the model’s spatial filters, especially at the premolar and the anterior regions. Anatomical uncertainty between the central and the lateral incisors was the cause of error, compounded where wear flattening or incisor crowding had occurred. Posterior errors were often compounded by intraoral mirror fogging at the distal areas, adding diffuse blur and hiding important morphological features such as cusp tips and fissures. This optical interference, especially prevalent where long exposure times were used to record the data, or with the patient having low breath control, significantly impaired the model’s capability to differentiate adjacent molars. The complication was further exaggerated where visual clutter came from orthodontic appliances and occlusal surfaces distorted through extensive restorative treatment or intensive wear.

A basic weakness of the pipeline is its inability to handle fully edentulous arches or unilateral edentulism since the estimation of missing teeth is based on interpolation between at least one anatomically valid tooth per side. Without bilateral anchors—such as the case where the entire quadrant is edentulous—x-center calculations break down, and spline-based placement is unable to provide robust estimations. In addition, the pipeline’s reliance on high-quality FDI assignment causes cascading weak points: if an existing tooth is mislabeled (e.g., tooth 15 predicted to be 14), the system may deduce an unfounded gap between teeth 13 and 14, even where anatomical continuity applies. Conversely, whenever several adjacent teeth are mislabeled, the resulting misalignment is able to generate non-existent edentulous areas or disregard true gaps.

These problems worsen with the pipeline’s module-by-module processing sequence, where the output of each module becomes the input of the next. An error in the initial modules, such as incorrect labeling of the jaw, bounding box incompleteness, or low-confidence number prediction, has the ability to irreparably damage downstream module output. Inadequate cross-module feedback prevents the system from re-examining or reversing previously made assumptions, thus leading to cumulative error propagation. All of these challenges point to the need for architectural enhancements, such as uncertainty-aware fusion between modules, dynamic error correction systems, and robustness training on uncommon or pathologic dental patterns.

The majority of published research on automated dental diagnostics has focused on radiography data, particularly panoramic views, that are gathered in strictly standardized settings with often little clinical variation. These models typically operate on well preprocessed datasets that lack the occlusal asymmetries, visual noise, and anatomical variability present in actual intraoral environments. Furthermore, a lot of earlier methods use single-stage architectures that make it difficult to understand anatomical reasoning, restrict the capacity to identify errors, and provide limited adaptability to complex or partial dentitions. Because of these drawbacks, they are consequently inappropriate for use in routine clinical practice. However, our pipeline is designed to function with regular intraoral pictures, overcoming common challenges such as saliva-induced glare, orthodontic artifacts, and partial dentition. Even under difficult imaging circumstances, our approach offers strong interpretability, modular scalability, and significant clinical output by breaking the problem down into anatomically grounded submodules. This presents our approach not as a minor enhancement but as a conceptual shift toward clinically deployable AI in dentistry.

Subsequent versions of the proposed pipeline may focus on moving from a function-dependent design to a graph-based anatomical reasoning framework. By integrating geometric deep learning methods such as Graph Neural Networks (GNN), the system could learn topological representations of the dental arch and perform context-aware analysis even in fully or partially edentulous cases—where current x-center–based heuristics fail due to the absence of adjacent teeth. An improvement of this kind would enable the model to deduce realistic anatomical configurations without depending on direct visual continuity, allowing for reliable missing tooth detection in increasingly intricate clinical situations. Furthermore, integrating the current pipeline with other specialized AI modules—for example, for implant planning, restoration quality assessment, caries detection, or gingival health evaluation—could turn it into a complete chairside diagnostic platform [31]. The smooth integration of this modular design into clinical procedures would facilitate real-time decision-making in a variety of dental specialties.

### 4.1 Risk of bias

In spite of demonstrating efforts to standardize data collection and imaging protocols, we acknowledge that a variety of potential biases may impact the results of this study. First, the recruitment of patients from only private dental clinics introduces a selection bias and affects generalizability of the model to patients that may also present in a public healthcare situation or in low-income communities.

In addition, excluding third molars due to their low prevalence potentially decreases anatomical diversity in the dataset, and compromises the model’s ability to generalize to atypical dental presentations. Even though this decision was made so that a balance could be achieved in the dataset; in order to prevent instability in the distribution of images upon training, it introduces a certain type of dataset bias that is important to recognize [32].

Furthermore, while manual preprocessing steps such as image cropping and implementation of quality control procedures were standardized, variability due to the operator does still exist and must be taken into account during data collection. Different annotators may interpret and implement annotation protocols differently and if strict accuracy is not enforced across the annotators, label accuracy will be impacted and impact model performance [33].

Finally, although the photographs were taken in controlled clinical conditions, the nature of intraoral imaging allows for variability in angulation, lighting, soft tissue interaction and artifacts. These noted factors will also have an adverse effect on feature extraction, which should be taken into consideration upon interpreting model outputs and implementation of the system.

## 5. Conclusions

Our study successfully developed a multi-stage AI pipeline for detecting and localizing bounded edentulous spaces from intraoral occlusal photographs. Despite the variability of real-world clinical imaging, the system demonstrated reliable diagnostic performance, underscoring the potential of AI-driven tools to support routine dental assessments. These outcomes align with our research objectives and contribute to the broader vision of advancing artificial intelligence to the next level of integration within dental diagnostics.

## Data Availability

All data produced in the present study are available upon reasonable request to the authors

## Credit authorship contribution statement

**Ehsan Shirdel:** Conceptualization, Methodology, Software, Formal analysis, Visualization, Writing – original draft, Resources, Writing – review and editing, Data curation.

**Saeideh Azizipour:** Writing – review and editing, Data curation, Methodology, Visualization.

**Mary Magdalyanova:** Validation, Writing – review and editing, Supervision.

## Declaration of competing interest

The authors declare that they have no known competing financial interests or personal relationships that could have appeared to influence the work reported in this paper.

## Declaration of generative AI and AI-assisted technologies in the writing process

During the preparation of this work the authors used OpenAI’s ChatGPT in order to improve the readability and language of the manuscript. After using this tool, the authors reviewed and edited the content as needed and take full responsibility for the content of the published article.

## Funding statement

This research did not receive any specific grant from funding agencies in the public, commercial, or not-for-profit sectors.

## Availability of data and material

The datasets used and analyzed during the current study are available from the corresponding author upon reasonable request.

## Acknowledgments

The authors would like to express their sincere gratitude to Dr. Aslan Etminan, Dr. Parisa Pouya, Dr. Mahsa Pouya and Dr. Ali Akbar Ghorbani for their invaluable support and insightful guidance throughout the development of this study.

## Author note

The authors reserve the right to determine the final authorship of this work. Individuals who have contributed to this study are currently under consideration for inclusion as co-authors, and any changes to the author list will be finalized prior to journal submission. This is a preliminary version and subject to revision before formal publication.

## Notes

### Competing Interest Statement

The authors have declared no competing interest.

### Funding Statement

This study did not receive any funding

### Author Declarations

Ethics Committee/IRB of I.M. Sechenov First Moscow State Medical University gave ethical approval for this work.

